# Who is (Not) Complying with the Social Distancing Directive and Why? Testing a General Framework of Compliance with Multiple Measures of Social Distancing

**DOI:** 10.1101/2020.10.26.20219634

**Authors:** Russell H. Fazio, Benjamin C. Ruisch, Courtney A. Moore, Javier A. Granados Samayoa, Shelby T. Boggs, Jesse T. Ladanyi

## Abstract

A study involving over 2000 online participants tested a general framework regarding compliance with a directive in the context of the COVID-19 pandemic. The study featured not only a self-report measure of social distancing but also behavioral measures -- simulations that presented participants with graphical depictions mirroring multiple real-world scenarios and asked them to position themselves in relation to others in the scene. The conceptual framework highlights three essential components of a directive: (1) the source, some entity is advocating for a behavioral change; (2) the surrounding context, the directive is in response to some challenge; and (3) the target, the persons to whom the directive is addressed. Belief systems relevant to each of these three components are predicted, and were found, to relate to compliance with the social distancing directive. The implications of the findings for public service campaigns encouraging people to engage in social distancing are discussed.

## Introduction

Given the current lack of a vaccine, minimizing the spread of COVID-19 requires that people change their behavior. People are urged to wash their hands frequently, use hand sanitizer, disinfect surfaces, and wear masks. Above all, government leaders and health experts plead with citizens to engage in social distancing – that is, deliberately increase the physical space between themselves and other people. The mantra “stay six feet away from others” has been repeated regularly.

Rarely has the entire population been called upon to exhibit immediate behavior change in compliance with an urgent directive. That raises an important question: who is or is not complying? Understanding who chooses to practice (or not) social distancing – and why – is crucial for the design of effective public service campaigns, both now, and during the occurrence of future pandemics. Whom should such campaigns target? What specific beliefs should be addressed?

Theory and research concerning compliance (behavioral change in response to an explicit or implicit request) is central to social psychology. The field has acquired substantial knowledge regarding social influence tactics promoting compliance [1], including such classic approaches as the foot-in-the-door [2], door-in-the-face [3], and low-balling [4]. In addition, the impact of both descriptive and injunctive norms has been examined extensively [5, 6].

However, as some scholars have noted [7], the field lacks a general theoretical framework about who is likely to comply with a directive, and why they might or might not. Such a framework is particularly important when considering a directive calling for compliance and behavior change on a large scale, as is currently true of the social distancing directive. The major aim of the current research is to test such a theoretical framework regarding the who and why of compliance.

## Compliance with a Directive: What’s Involved?

Any directive is open to interpretation and ultimately will be assessed as warranting or not warranting compliance. Only if deemed justified is the directive likely to promote behavioral change. Yet, one of the core principles of social psychology is that individuals construct their own reality [8-10]. Such constructions influence and are influenced by the information to which individuals choose to expose themselves [11], their exploratory behavior, and ultimately the accuracy of their understanding of reality [12, 13]. Thus, any given directive will be viewed through the lens of the individual’s knowledge, beliefs, and attitudes. Decades of research demonstrate the pervasive influence of such factors on judgments and decisions [14-16].

Such considerations raise the question of what might be regarded as the essential components of a directive. Our theoretical framework highlights three: (1) the *source:* the entity advocating for behavioral change; (2) the surrounding *context:* the challenge the directive addresses; and (3) the *target:* the persons to whom the directive is addressed. Is the source to be trusted? What does the surrounding context imply about the seriousness of the challenge? Are there individual propensities that affect responsivity to the directive? Thus, a complex network of beliefs will affect who chooses to comply or not, and for what reasons. Some individuals’ belief systems will lead them to assess the directive favorably, thus promoting behavior change. Others will reach less positive conclusions and, hence, fail to respond appropriately.

## Measuring Social Distancing

In examining compliance, the challenge rests in how to assess social distancing. The field’s dominant approach is to ask people to report the frequency with which they socially distance. However, the problems associated with self-reports of behavior have been discussed for decades. Individuals may over-report their social distancing to convey a socially desirable impression to others and themselves [17-20]. Moreover, self-reports may be all the more problematic to the extent that they rely on retrospective memory regarding past behavior [21, 22]. Even more troubling, some of the very characteristics and beliefs we predict will affect responsiveness to the directive may also influence how a person (mis)represents their social distancing on self-report measures.

We thus supplemented self-reports of social distancing with an innovative, more behaviorally-oriented approach. We simulate social distancing behavior with graphical depictions mirroring real-world scenarios, asking participants to position themselves in relation to others in the scene. For example, in one scenario people view a street scene and choose whether to follow a path passing directly in front of another individual, or detour via the crosswalk. Other more interactive measures allow participants to manipulate a slider to virtually “distance” themselves from an oncoming walker, or to separate individuals waiting in line. These graphical scenarios simulate real-life and require a concrete, “in-the-moment” behavioral decision. In this sense these measures are behavioral in nature.

Our argument regarding the value of these behavioral scenarios parallels a relevant empirically-supported proposition regarding attitudes as predictors of behavior. Attitude measures are more likely to predict behavior when they match the behavior in terms of specificity regarding the action in question and the context in which the action is performed [23, 24]. Similarly, the simulated scenarios closely match real-life situations in terms of their concreteness. They offer a means, in addition to a self-report, of indexing the extent to which individuals make decisions that accord with the principle of social distancing.

## Predictor Variables

This study examines the relation between social distancing and various predictor variables. To test our guiding framework regarding compliance, the predictors involve our three classes of beliefs – those regarding the source of the directive, the surrounding context posed by the challenge, and additional characteristic of the targets themselves.

### Beliefs about the source

The primary source of the social-distancing directive is government and health officials. The latter are medical scientists or liaisons represent the scientific community. Given the distinction in the literature between valuing science as means of acquiring knowledge and trusting scientists and their work [25], we hypothesized that (a) greater belief in science and (b) greater trust in scientists would relate positively to compliance.

Given the highly polarized sociopolitical context and the politicization of the pandemic, assessing faith in government officials is more complex. President Trump downplayed the pandemic’s severity relative to state Governors. Accordingly, we separately assessed trust in the President’s and Governors’ leadership regarding the pandemic, predicting that these measures may relate differently to compliance.

### Beliefs about the context

Several questions regarding assessments of the pandemic and support for social distancing were included to examine the hypothesis that greater concern about the virus and positive attitudes toward the directive would be associated with more social distancing. We tested a similar hypothesis regarding accurate knowledge about COVID-19 by administering a brief quiz about the virus. More knowledgeable individuals were expected to display more distancing.

### Target Characteristics

Two sets of target characteristics were expected to relate to individuals’ receptivity to the plea to engage in social distancing: beliefs relevant to disease or views of the government and more general characteristics of the individual. Perceived vulnerability to disease [26] and its concomitant disgust sensitivity [27] were expected to relate positively to social distancing. Similarly, general compassion [28] and concern for others’ vulnerability to the coronavirus were expected to predict distancing. Given President Trump’s equivocal stance and the traditional emphasis that conservatives place on economic freedom, we anticipated that conservatism would be associated with less distancing. In addition, we predicted that conspiratorial ideation [29, 30] would promote minimization of COVID-19’s severity, and hence relate to less compliance.

Turning to the second set of target characteristics, we also predicted that individual differences in scientific literacy [31] would likely relate to both trust in health experts and the development of accurate knowledge regarding the coronavirus. Hence, scientific literacy was expected to be associated with more distancing. Additionally, a considerable literature highlights the importance of the particular news sources that individuals follow [32, 33]. We expected that reliance on more conservative news sources would relate to minimizing the threat posed by the pandemic and less distancing behavior.

## Materials and methods

We recruited a sample from Mechanical Turk. Although not representative of the U.S. population, MTurk samples are considerably more diverse than the student samples used in most psychological research [34, 35], and they perform similarly to non-MTurk samples across many tasks and measures [36, 37], including surveys on political attitudes [38]. Further, our aim is not to make claims regarding the absolute frequency of beliefs and behaviors in the population, but rather to understand how the psychological variables of interest relate to social distancing behavior. Given these aims, we judged the MTurk sample as appropriate for testing our hypotheses. As will become evident, both the very systematic nature of the data and their replication of some relations previously established in the literature attest further to the appropriateness of the MTurk sample.

Past experience with MTurkers led us to believe that they prefer, and respond most conscientiously, when a study is relatively short. Hence, we included only our social distancing measures, survey items assessing beliefs and knowledge about the pandemic, and various demographics as the elements of a common survey that was completed by all the participants. Subsets of our other predictors were included in four distinct surveys to which participants were randomly assigned. The four subsets involved: (a) source beliefs and science literacy, (b) news sources and belief in conspiracy theories, (c) compassion and concern for others vulnerability to COVID-19, and (d) perceived vulnerability to disease and disgust sensitivity.

### Participants

We aimed for sample sizes that would clearly be large enough to obtain stable estimates of the relations with social distancing within each of our four sub-studies [39]. A total of 2,004 MTurkers (US residents) participated in the common survey (903 women, 1,084 men, 17 no response; *M*_age_ = 38.66, *SD*_*age*_ = 12.33), with about 500 being randomly assigned to each sub-study. They completed the study on May 7-8, 2020, at which time some states had begun to re-open their economies.

### Measures

Ohio State University’s Institutional Review Board approved all study procedures (IRB: 2020B0129). After providing informed consent, participants completed the behavioral measures of social distancing, followed by questions regarding the pandemic, the test of COVID knowledge, the unique set of predictor variables for the study to which the participant had been randomly assigned, and finally a series of demographic questions. All of the measures and the datafile are available at https://osf.io/359et/?view_only=2c83bae138984cb592ba354043b3d1dc.

### Social Distancing Behaviors

Ten graphic scenarios comprised the behavioral measure of social distancing. Examples include: (a) An image of two people approaching each other in a crosswalk. Participants moved a slider that shifted the walkers from the center of the crosswalk to the distance that they would prefer. (b) An aerial image of a crowded beach for which participants were asked to “click the point on the beach where you’d be most likely to lay down your towel.” The distance to the nearest person (in pixels) served as the measure of interest. (c) A graphic depicting a park for which participants used a 4-point scale to indicate whether would they definitely or probably walk via one of two paths. One path was less direct, but also more isolated relative to the many people situated on either side of the alternative path. All ten of the behavioral scenarios are described in the Supplemental Material and can be viewed at our demonstration website, http://psychvault.org/social-distancing-measures/. After standardizing scores from each measure, we computed the average as our index of social distancing behaviors (α = 82).

### Predictor Variables

#### Questions Regarding the Pandemic

The behavioral scenarios were followed by the common portion of the survey, including the self-report measure of social distancing: “Generally speaking, how strictly have you personally been following the “social distancing” recommendations?” and a number of questions regarding perceptions of the pandemic. Participants were asked how worried they were about contracting the virus, how likely they were to do so, and how concerned they were about the spread of the virus. They also indicated whether they felt the threat of COVID-19 had been “greatly exaggerated” versus “not conveyed strongly enough” and whether economic considerations or public safety should receive priority. A more general attitudinal question asked participants to indicate the extent to which they supported or opposed the guideline to engage in social distancing.

#### COVID-19 Knowledge

Participants indicated whether statements regarding the coronavirus (facts and myths addressed by the Centers for Disease Control and Prevention and the World Health Organization) were true or false. Included were false items such as “Antibiotics are an effective treatment for COVID-19 / the coronavirus” and true items such as “Some individuals who have COVID-19 / the coronavirus do not show any symptoms.” The number answered correctly served as our index of COVID knowledge (α =.83).

#### Faith in Government

To assess people’s trust in different elements of the government, we used four single-item measures. Specifically, participants were asked to rate the extent to which they “trust President Trump to lead us effectively through the current COVID-19 crisis” and separately whether they trust state governors to do so. They also indicated the extent of their general confidence in President Trump and general confidence that the federal government will address the nation’s problems effectively.

#### Belief in the Value of Science

To assess the extent to which individuals believe in the value of science as the best way to accumulate knowledge about the world, participants responded to a shortened version (the six items with the highest factor loadings) of a scale developed by Farias, Newheiser, Kahane, & de Toledo [40]. Participants rated the degree to which they endorsed statements such as “Science is the most efficient means of attaining truth” (α = .92).

#### Trust in Scientists

This variable was assessed using a shortened version (the 11 items, out of 21, with the highest corrected item-total correlations) of the scale developed by Nadelson et al. [25]. Participants rated their level of agreement with statements such as “We should trust the work of scientists” (α = .80).

#### Science Literacy

Participants’ understanding of basic scientific ideas was assessed using the Civic Scientific Literacy Scale [31]. This scale consists of 11 claims such as “Light travels faster than sound” and “Electrons are smaller than atoms” for which participants indicate agreement or disagreement (α = .59).

#### Conspiracy Theories

Participants completed the Generic Conspiracist Beliefs scale [29]. They rated each of 15 items that address a variety of generic conspiracy theories, e.g., “Evidence of alien contact is being concealed from the public” (α = .96).

#### News Sources

Participants were presented with a list of potential News sources: CNN, Fox News, MSNBC, NPR, national newspapers and magazines, social media, and ABC, CBS, or NBC News, as well as the option “do not follow the news.” They were asked to select all the sources from which they got their news in the past week. Any who did not indicate that they do not follow the news were then asked to select which one of the news outlets they consider to be their primary source of news.

#### Compassion

To assess general compassion for others, we employed a subset of items of the Interpersonal Reactivity Index [28]. Specifically, we included the 14 items of the scale related to empathic concern (e.g., “When I see someone being taken advantage of, I feel kind of protective towards them”) and perspective taking (e.g., “Before criticizing somebody, I try to imagine how I would feel if I were in their place”). Participants indicated the extent to which each statement described them (α = .87).

#### Concern for Others’ Vulnerability to COVID-19

Four items assessed the extent to which participants experienced empathic concern for people who had contracted COVID-19 or were vulnerable to do so. Participants rated their agreement with such statements as “I feel it is my personal responsibility to keep others safe from COVID-19 coronavirus” (α =.67).

#### Perceived Vulnerability to Disease

Individuals’ perceptions of their likelihood of contracting a disease or illness was assessed with the 15-item scale developed by Duncan, Schaller, & Park [26]. Participants rated the degree to which they agreed with statements such as “If an illness is ‘going around’ I will catch it” (α = .73).

#### Disgust Sensitivity

The contamination subscale (five items) from the Disgust Scale Revised [27] was used to assess individuals’ sensitivity regarding situations that have the potential for the transmission of pathogens. Participants rated how disgusted they would be by various scenarios as well as their agreement with statements such as “I probably would not go to my favorite restaurant if I found out the cook had a cold” (α = .70).

#### Demographics

In addition to a number of demographic questions, participants were asked to identify their political orientation on a scale ranging from 1 (Extremely liberal) to 7 (Extremely conservative).

#### Attention check

The survey concluded with a brief attention check in which participants were informed that a man had seen a beautiful butterfly, and were then asked to select what he had seen: a girl, a day, a fruit, or an insect. Ninety-one percent correctly chose insect. To provide a more conservative test of our hypotheses, we did not exclude participants who failed this attention check. However, none of our conclusions or statistical results are altered to any meaningful degree if these participants are excluded from analyses.

## Results

### Social Distancing

To test each of the hypotheses, we examined the multiple correlation between a given variable and our two indices of social distancing – the behavioral and the self-report measures. Table 1 presents the regression data for each of our hypothesized predictor variables. Table 2 displays the correlations among the variables. Turning first to the source, both *belief in the value of science* and *trust in scientists* correlated positively with social distancing. As expected, these variables also correlated with assessments of the pandemic itself, including more support for the social distancing guidelines, greater concern about the spread of COVID-19, stronger beliefs that the threat posed by the virus had not been exaggerated, and a view that public safety should be prioritized over economic recovery.

**Table 1.** Multiple Correlations with Behavioral and Self-Reported Social Distancing.

|  |  |  |  | Standardized Beta<sup>a</sup> |  |
| --- | --- | --- | --- | --- | --- |
|  | R | F | df | Behavioral | Self-Report |
| **Beliefs about the Source** |  |  |  |  |  |
| Belief in Value of Science | .247 | 15.993*** | 494 | .117* | .172*** |
| Trust in Scientists | .372 | 39.754*** | 494 | .299*** | .127** |
| Trust President Trump re COVID-19 crisis | .294 | 23.337*** | 493 | -.309*** | .038 |
| Trust State Governors re COVID-19 crisis | .268 | 19.021*** | 493 | .027 | .255*** |
| Confidence in Federal Gov’t Effectiveness | .205 | 10.832*** | 493 | -.223*** | .139** |
| General Confidence in President Trump | .281 | 21.118*** | 493 | -.294*** | .033 |
| **Beliefs about the Context** |  |  |  |  |  |
| Support Social Distancing Guideline | .650 | 728.207*** | 1995 | .248*** | .498*** |
| Worry about Contracting Virus | .303 | 101.249*** | 1998 | .145*** | .208*** |
| Likely to Contract Virus | .096 | 9.327*** | 1997 | .026 | .081** |
| Concerned about the Spread of the Virus | .409 | 200.380*** | 1997 | .221*** | .257*** |
| Threat (not) exaggerated | .458 | 265.539*** | 1998 | .343*** | .185*** |
| Economy More Important than Safety | .377 | 165.181*** | 1997 | -.162*** | -.274*** |
| COVID Knowledge | .286 | 89.128 | 1998 | .269*** | .034 |
| **Other Receptivity-Related Beliefs** |  |  |  |  |  |
| General interpersonal compassion | .359 | 36.753*** | 496 | .134** | .275*** |
| Concern for others’ COVID vulnerability | .501 | 83.228*** | 496 | .122** | .432*** |
| Disgust Sensitivity | .151 | 5.888** | 502 | -.020 | .159** |
| Perceived vulnerability to disease | .227 | 13.632*** | 502 | .116* | .150** |
| Political ideology (higher, more conservative) | .228 | 54.526*** | 1996 | -.183*** | -.075** |
| Belief in conspiracy theories | .257 | 17.522*** | 496 | -.249*** | -.016 |
| **Other Target Characteristics** |  |  |  |  |  |
| Age | .166 | 28.393*** | 1996 | .137*** | .051* |
| Gender (1=male/0=female) | .126 | 16.018*** | 1984 | -.107*** | -.034 |
| Science Literacy | .198 | 10.026*** | 494 | .178*** | .038 |
| Fox News<sup>b</sup> | .177 | 7.650** | 475 | -.190*** | .033 |
| NPR<sup>b</sup> | .118 | 3.325* | 475 | .079 | .058 |
| Papers, Magazines<sup>b</sup> | .141 | 4.790** | 475 | .093 | .071 |

<sup>a</sup> Higher numbers reflect more social distancing behavior.
<sup>b</sup> coded 0=neither watch last week, nor primary news source, 1=watched last week or primary, 2=both
\* $p < .05$ ; \*\* $p < .01$ ; \*\*\* $p < .001$

**Table 2.** Correlation Matrix.

|  | A | B | C | D | E | F | G | H | I | J | K | L | M | N | O | P | Q | R | S | T | U | V | W | X | Y | Z | AA |
| --- | --- | --- | --- | --- | --- | --- | --- | --- | --- | --- | --- | --- | --- | --- | --- | --- | --- | --- | --- | --- | --- | --- | --- | --- | --- | --- | --- |
| A. Belief in Value of Science |  |  |  |  |  |  |  |  |  |  |  |  |  |  |  |  |  |  |  |  |  |  |  |  |  |  |  |
| B. Trust in Scientists | .289** |  |  |  |  |  |  |  |  |  |  |  |  |  |  |  |  |  |  |  |  |  |  |  |  |  |  |
| C. Trust Trump re COVID-19 | -.105* | -.528** |  |  |  |  |  |  |  |  |  |  |  |  |  |  |  |  |  |  |  |  |  |  |  |  |  |
| D. Trust Governors re COVID-19 | .185** | .007 | .311** |  |  |  |  |  |  |  |  |  |  |  |  |  |  |  |  |  |  |  |  |  |  |  |  |
| E. Confidence in Federal Gov't | .040 | -.395** | .767** | .423** |  |  |  |  |  |  |  |  |  |  |  |  |  |  |  |  |  |  |  |  |  |  |  |
| F. General Confidence in Trump | -.101* | -.536** | .927** | .289** | .760** |  |  |  |  |  |  |  |  |  |  |  |  |  |  |  |  |  |  |  |  |  |  |
| G. Support Social Distancing | .342** | .366** | -.195** | .364** | -.016 | -.164** |  |  |  |  |  |  |  |  |  |  |  |  |  |  |  |  |  |  |  |  |  |
| H. Worry Contracting Virus | .256** | -.113* | .040 | .180** | .124** | .071 | .330** |  |  |  |  |  |  |  |  |  |  |  |  |  |  |  |  |  |  |  |  |
| I. Likely to Contract Virus | .169** | -.232** | .074 | .138** | .099** | .093** | .148** | .673** |  |  |  |  |  |  |  |  |  |  |  |  |  |  |  |  |  |  |  |
| J. Concern about Spread of Virus | .198** | .212** | -.098* | .226** | -.066 | -.107* | .396** | .387** | .205** |  |  |  |  |  |  |  |  |  |  |  |  |  |  |  |  |  |  |
| K. Threat (not) exaggerated | .113* | .587** | -.495** | .024 | -.374** | -.528** | .456** | .082** | -.054* | .290** |  |  |  |  |  |  |  |  |  |  |  |  |  |  |  |  |  |
| L. Economy versus Safety | -.209** | -.217** | .286** | -.134** | .122** | .251** | -.479** | -.320** | -.191** | -.267** | -.338** |  |  |  |  |  |  |  |  |  |  |  |  |  |  |  |  |
| M. COVID Knowledge | -.007 | .644** | -.464** | -.141** | -.426** | -.487** | .154** | -.300** | -.368** | .083** | .547** | -.019 |  |  |  |  |  |  |  |  |  |  |  |  |  |  |  |
| N. Interpersonal compassion |  |  |  |  |  |  | .308** | -.029 | -.087 | .170** | .327** | -.171** | .370** |  |  |  |  |  |  |  |  |  |  |  |  |  |  |
| O. Concern for others |  |  |  |  |  |  | .481** | .307** | .191** | .274** | .221** | -.259** | .086 | .483** |  |  |  |  |  |  |  |  |  |  |  |  |  |
| P. Disgust Sensitivity |  |  |  |  |  |  | .159** | .347** | .244** | .127** | -.175** | -.069 | -.390** |  |  |  |  |  |  |  |  |  |  |  |  |  |  |
| Q. Vulnerability to disease |  |  |  |  |  |  | .213** | .335** | .225** | .190** | .119** | -.108* | .010 |  |  | .356** |  |  |  |  |  |  |  |  |  |  |  |
| R. Politically conservative | -.300** | -.375** | .522** | .045 | .376** | .522** | -.233** | -.053* | -.011 | -.144** | -.388** | .284** | -.276** | -.184** | -.171** | .170** | -.043 |  |  |  |  |  |  |  |  |  |  |
| S. Belief in conspiracy theories |  |  |  |  |  |  | -.136** | .186** | .260** | -.002 | -.376** | .035 | -.571** |  |  |  |  | .128** |  |  |  |  |  |  |  |  |  |
| T. Age | -.082 | .107* | -.007 | .039 | -.074 | .017 | .055* | -.075** | -.131** | .096** | .069** | .046* | .164** | .127** | -.028 | -.036 | -.110** | .097** | -.218** |  |  |  |  |  |  |  |  |
| U. Gender (1=male/0=female) | .141** | -.119** | .091* | .102* | .134** | .088 | -.056* | -.018 | .041 | -.086** | -.140** | -.007 | -.182** | -.221** | -.167** | -.009 | -.087 | .036 | .084 | -.117** |  |  |  |  |  |  |  |
| V. Area Re-opened (1=Yes/0=No) | .054 | -.124** | .120** | -.048 | .141** | .120** | -.044 | .049* | .116** | -.015 | -.131** | .047* | -.204** | -.050 | -.026 | .115** | .041 | .093** | .228** | -.007 | .022 |  |  |  |  |  |  |
| W. Science Literacy | .007 | .514** | -.396** | -.125** | -.436** | -.409** | .021 | -.260** | -.267** | .030 | .348** | .089* | .621** |  |  |  |  | -.211** |  | .212** | -.079 | -.170** |  |  |  |  |  |
| X. Fox News<sup>b</sup> |  |  |  |  |  |  | -.168** | -.032 | -.015 | -.053 | -.275** | .191** | -.149** |  |  |  |  | .412** | .151** | .126** | -.007 | .112* |  |  |  |  |  |
| Y. NPR<sup>b</sup> |  |  |  |  |  |  | .157** | .000 | .090* | .071 | .162** | -.122** | .114* |  |  |  |  | -.221** | -.032 | -.025 | .057 | -.087 |  | -.196** |  |  |  |
| Z. Papers, Magazines<sup>b</sup> |  |  |  |  |  |  | .124** | .006 | .047 | .106* | .167** | -.053 | .210** |  |  |  |  | -.210** | -.220** | .035 | -.007 | -.092* |  | -.191** | .124** |  |  |
| AA. Self-Report Distancing | .223** | .257** | -.096* | .267** | .042 | -.095* | .611** | .275** | .093** | .359** | .342** | -.348** | .158** | .339** | .490** | .150** | .202** | -.159** | -.137** | .114** | -.083** | -.052* | .115* | -.057 | .095* | .114* |  |
| BB. Behavioral Distancing | .192** | .354** | -.292** | .138** | -.162** | -.279** | .476** | .241** | .064** | .339** | .428** | -.287** | .284** | .266** | .328** | .051 | .183** | -.218** | -.256** | .160** | -.122** | -.095** | .195** | -.174** | .106* | .126** | .459** |

\*\* $p < .01$ ; \* $p < .05$ ; Cells without an entry involve variables that were included in different sub-studies.

*Faith in the government officials* was more complex, just as we anticipated. Whereas greater trust that the State Governors can lead us effectively through the COVID-19 crisis was positively associated with behavioral compliance, the relations were negative when participants considered either President Trump specifically or the federal government more generally. More confidence in those sources was associated with *less* social distancing. Interestingly, these relations do not appear to be a simple reflection of political orientation. Although participants who more strongly identified as conservative engaged in less social distancing and expressed more trust/confidence in President Trump, in each case the measures of social distancing accounted for unique variance over and above that explained by political orientation (all *p*’s < .001).

All the *belief measures concerning the pandemic itself* related as expected with social distancing. This was especially true of support for the social distancing guideline, worry about contracting the virus, concern about the spread of the virus, and the assessment that the threat posed by the virus had not been exaggerated. Believing that relatively more emphasis should be placed on economic recovery than public safety also was associated with less social distancing.

Answers to our test of *COVID-19 knowledge* also related positively to behavioral compliance. Importantly, the recognition of true statements and the rejection of misinformation each correlated with social distancing (multiple *R*’s of .234 and .260, respectively). Knowledge also was associated with support for the social distancing guideline and especially with the belief that the threat posed by the coronavirus had not been exaggerated. In addition, more knowledgeable individuals expressed greater trust in scientists and less confidence in President Trump.

Self-beliefs highlighting *interpersonal compassion* and *concern for others’ vulnerability* to the virus were associated with more social distancing. These variables correlated as expected with beliefs about the pandemic. For example, more compassionate individuals were more supportive of the social distancing guideline and believed that the threat of the virus had not been exaggerated. The same was true of individuals who had expressed concern for others’ vulnerability; they also were more worried that they themselves would contract the virus.

The data offered a number of interesting observations regarding the extent to which respondents viewed themselves as generally *vulnerable to disease*. This variable was related to more social distancing, and, just as one would expect, with greater worry about contracting COVID-19 and greater perceived likelihood of contracting it. Disgust sensitivity correlated with perceived vulnerability to disease, replicating past findings, and also related to social distancing. Stronger disgust sensitivity also was associated with greater worry about contracting COVID-19 and greater likelihood of doing so.

As already noted, *political orientation* also was relevant; more conservative individuals engaged in less physical distancing. Just as expected, political ideology correlated strongly with general confidence in President Trump and trust in his leadership regarding the COVID-19 crisis, but not with trust in the state Governors. More conservative individuals also reported less belief in the value of science and less trust in scientists. They also believed the threat of the coronavirus to have been exaggerated and that economic considerations needed to take priority over public safety.

Generally believing in *conspiracy theories* also was predictive of less social distancing, possibly because it promotes a less accurate view of the pandemic. Indeed, such beliefs correlated strongly with scores on the test of COVID-19 knowledge. Conspiracy theorists also were more likely to believe that the threat posed by the coronavirus had been exaggerated.

In addition to beliefs, we examined a number of other personal characteristics that seemed potentially relevant to receptivity to the directive (see the fourth section of Table 1). Female participants displayed more evidence of social distancing, as did older participants. Our hypothesis regarding *science literacy* also received support. Those who exhibited a greater understanding of a small set of basic scientific facts engaged in more social distancing. Scientific literacy also related strongly to expressed trust in scientists and scores on the test of COVID-19 knowledge. It also was associated with the belief that the threat posed by the virus had not been exaggerated.

Finally, although none of the multiple correlations were very substantial, accounting for less than 3% of the variance, a number of the *news sources* variables related to social distancing. Whereas engagement with NPR or newspapers and magazines was associated positively social distancing, greater involvement with Fox News related negatively to distancing. The latter was more common for participants who endorsed a more conservative political orientation, whereas the former was associated more strongly with a more liberal perspective. Parallel relations were observed with support for the social distancing guideline, valuing economic considerations more than public safety, believing that the COVID-19 threat had been exaggerated, and accurate knowledge regarding COVID.

## Comparing Behavioral and Self-Reported Social Distancing

Our primary interest was to employ the behavioral and self-report measures of social distancing as supplemental to one another and, hence, as jointly related to each of the hypothesized predictor variables. Although the two measures were related (*r* = .459, *p* < .001), the magnitude of the correlation was not so overwhelming as to suggest that they were equivalent. Given this observation, it is interesting to consider how the two variables differ with respect to the unique variance for which they each accounted in the multiple regressions (Table 1). Although drawing any strong inferences from the patterns evident in the table is difficult, the larger differences appear especially striking. For COVID knowledge and scientific literacy, which are much more objective measures than any of the other predictor variables, the multiple correlation was driven almost entirely by the behavioral measure. Indeed, the self-report failed to account for any significant unique variance. The same was true with respect to engagement with Fox News, belief in conspiracy theories, and the variables reflecting trust in President Trump’s leadership regarding the pandemic and general confidence in him. On the other hand, for the arguably most subjective measure – support or opposition for the social distancing guideline – the self-report measure of social distancing accounted for twice the unique variance that was associated with the behavioral measure. Similar patterns were evident for general compassion, concern for others’ vulnerability to COVID-19, and disgust sensitivity. These differences certainly suggest that behavioral and self-report measures of social distancing are not responsive to the same forces.

## Discussion

The findings highlight the importance of individuals’ beliefs as factors associated with social distancing behavior. They also support the theoretical framework of compliance that guided our selection of variables for inclusion in the study. Any directive regarding behavior change will be shaped by beliefs about the directive’s source, beliefs about the context surrounding the challenge to which the directive is responding, and relevant self-views and characteristics. As such, the framework is applicable to any call for behavior change aimed at the general public. When applied to the social-distancing directive, the conceptual framework led to our focus on (a) source variables related to the government and public health officials, (b) beliefs regarding COVID-19 and the severity of the threat, and (c) various self-related beliefs and target characteristics influencing receptivity to the social distancing directive.

Importantly, these relations were evident not only on a self-report social-distancing measure but also on a measure that relied on vivid, graphical simulations of real-life behavior. Participants made concrete, “in-the-moment” decisions about actions involving different degrees of social distancing. They interactively distanced themselves from oncoming passersby and from people standing in line. They selected a position on a crowded beach. As such, the behavioral decisions closely matched the features of real-life situations.

The current findings did indeed reveal some striking differences between behavioral and self-report measures of social distancing. Although the two were related, the correlation did not reach a level that suggested these were equivalent measures of the same construct. Moreover, the unique variance accounted for by each measure differed markedly for a number of predictor variables. Especially telling was that scores on our tests of COVID knowledge and scientific literacy – the most objective of our predictor variables – related more strongly to the behavioral measures, with self-reports accounting for little or no additional variance. Thus, self-reports do not cohere with behavioral decisions sufficiently to suggest they are mutually interchangeable. However, they complement one another well, as is evident by their accounting for unique variance for many predictor variables.

We conclude with a brief consideration of the implications of the present findings for public service campaigns encouraging social distancing. How might compliance be promoted? The findings highlight the importance of communicating accurate knowledge and dispelling misinformation. Campaigns also should promote science as the means by which we will come to understand the coronavirus, how it spreads, and how it might be contained. There is also likely value in appealing to, and heightening concern, about others’ vulnerability to the coronavirus and the suffering of those infected. Similarly, the results regarding perceived vulnerability to disease and compassion suggest the need to emphasize the vulnerability of people of all ages to the virus and the role that everyone, whether symptomatic or not, plays in spreading it. Indeed, the data seem to call for frequent repetition of the portrayal of social distancing guidelines that White House coronavirus response coordinator Dr. Deborah Birx offered: “This is a road map to prevent your grandmother from getting sick” [41].

Maybe, most importantly, the findings highlight the importance of the plea so pointedly articulated by World Health Organization Director-General Ghebreyesus: “Please don’t politicize this virus” [42]. The politicizing is very evident in the present data. Very different relations were observed with respect to trust in President Trump versus the State Governors as providing effective leadership during the COVID-19 crisis. Participants’ political orientation, and even their exposure to more partisan news sources, related to beliefs about the severity of the CIVOD-19 threat, support for the social distancing guideline, and social distancing behavior. Obviously, a more coherent, non-partisan perspective, emphasizing reliance on scientific evidence, is essential.

## Data Availability

All of the measures and the datafile are available on the Open Science Framework at https://osf.io/359et/?view_only=2c83bae138984cb592ba354043b3d1dc.

https://osf.io/359et/?view_only=2c83bae138984cb592ba354043b3d1dc

## Supporting information

**S1 Supplemental Material**. Description of the Simulated Behavioral Measures of Social Distancing.

