## Supplemental Material for "Who is (Not) Complying with the Social Distancing Directive and Why? Testing a General Framework of Compliance with Multiple Measures of Social Distancing"

Description of the Simulated Behavioral Measures of Social Distancing

As noted in the text, the behavioral measure of social distancing consisted of ten graphical scenarios, each of which required a decision that could vary in its extent of social distancing. Briefly, they involved: (a) An image of two people walking along a wide path through the woods for which participants were asked to imagine being out for a walk with a friend. They moved a slider (scored as 0-10) that separated the two individuals to indicate the minimum distance they would feel comfortable having between the two of them. (b) A scene of a crowded grocery with ten figures representative of people for which participants moved a slider to "remove" people until they had reached the maximum number of people they would feel comfortable having in the grocery store with them. (c) An aerial image of a crowded beach for which participants were asked to "click the point on the beach where you'd be most likely to lay down your towel." The distance to the nearest person (in pixels) served as the measure of interest. (d) An image of two people approaching each other in the middle of a crosswalk for which participants were asked to imagine walking across the street. They moved a slider (scored as 0-10) that shifted the walkers from the center of the crosswalk to the distance that they would leave between the two of them. (e) An aerial image of a crowded plaza for which participants were to imagine that they needed to traverse the plaza by drawing a path from a start point located the southwest end of the plaza to an end point at the northeast end. The length of the paths that participants drew (in pixels) was measured as the data of interest. (f) A scene of people standing in line at a bus stop for which participants moved a slider (scored as 0-10) that separated the people to the distance they would "want between you and the other people" in line. (g) Each of the final four scenarios asked participants to choose between two alternatives using a 4-point scale designating definitely or probably choosing option A or option B, one of which was the more socially distant alternative. The first of these graphical images presented a street scene and asked participants to choose whether to follow a path that would cross directly in front of another individual or to detour across the street via a crosswalk. (h) A graphic depicting a park that participants were asked to imagine they needed to walk through to get to a store on the other side. One path was less direct, but also much more isolated, in contrast to the many people situated on either side of the alternative path. (i) A depiction of a coffeeshop with two open seats at either side of the shop, one of which involved more social distancing because the rectangular table of four seats was larger than the alternative square table for four people. (j) A diagram of two equidistant routes to a library's book return desk along either side of dividing bookshelves, one of which had more people standing nearby than the other.[1] (The behavioral scenarios can be viewed at our demonstration website, http://psychvault.org/social-distancing-measures/.) After standardizing scores from each measure, we computed the average as our composite index of social distancing behaviors ($\alpha = 0.82$).

[1] Given that many states had not yet re-opened businesses, the coffeeshop and library items were preceded by a question that asked whether the particular area in which the participant lived had relaxed restrictions and allowed restaurants, stores, libraries, playgrounds, and the like to reopen. For the 60.1% who answered no, additional text instructed them to "imagine a future time when the pandemic has subsided sufficiently so that restaurants, stores, libraries, and playgrounds are open, but health officials continue to recommend that people engage in social distancing" while responding to the following scenarios.
